# Wavelet-Based Compression Method for Scale-Preserving SWIR Hyperspectral Data

**DOI:** 10.1101/2025.01.18.25320781

**Authors:** Hridoy Biswas, Rui Tang, Shamim Mollah, Mikhail Y. Berezin

## Abstract

**Purpose:** Hyperspectral imaging (HSI) collects detailed spectral information across hundreds of narrow bands, providing valuable datasets for applications such as medical diagnostics. However, the large size of HSI datasets, often exceeding several gigabytes, creates significant challenges in data transmission, storage, and processing. This study aims to develop a wavelet-based compression method that addresses these challenges while preserving the integrity and quality of spectral information.

**Approach:** The proposed method applies wavelet transforms to the spectral dimension of hyperspectral data in three steps: 1) wavelet transformation for dimensionality reduction, 2) spectral cropping to eliminate low-intensity bands, and 3) scale matching to maintain accurate wavelength mapping. Daubechies wavelets were used to achieve up to 32x compression while ensuring spectral fidelity and spatial feature retention.

**Results:** The wavelet-based method achieved up to 32x compression, corresponding to a 96.88% reduction in data size without significant loss of important data. Unlike PCA and ICA, the method preserved the original wavelength scale, enabling straightforward spectral interpretation. Additionally, the compressed data exhibited minimal loss in spatial features, with improvements in contrast and noise reduction compared to spectral binning.

**Conclusions:** This study demonstrates that wavelet-based compression is an effective solution for managing large HSI datasets in medical imaging. The method preserves critical spectral and spatial information, and therefore facilitates efficient data storage and processing, providing the way for practical integration of HSI technology in clinical applications.

## 1. INTRODUCTION

In the last two decades, hyperspectral imaging (HSI) has emerged as a promising technology in various fields, including medical diagnostics,^1^ environmental monitoring,^2^ agriculture,^3^ and mineralogy.^4^ By acquiring images across a broad range of wavelengths from UV to SWIR and beyond, HSI generates a 3D dataset that enables a large number of algorithms^5-7^ to extract specific features of objects and provide higher spatial contrast then traditional single wavelength or RGB based images. Despite the advantages of HSI, the sheer volume of hyperspectral data significantly strains storage capacities and computational resources, mandating effective data compression and noise reduction strategies. In contrast to a single band or typical color images that have relatively small file sizes (under 10 MB), hyperspectral images capture data from a much larger number of spectral bands and covering larger electromagnetic spectrum. Depending on the instrumentation, modern HSI systems (bench-type, field deployed, remote sensing through satellites and drones) collect data across dozens or hundreds of spectral bands where the file size for a typical for example bench-type HS imager can reach a gigabyte or more for a single dataset. Such problem is particularly critical for real-time applications and onboard processing in remote sensing and medical imaging. The large size of HSI datasets overwhelm memory capacity and computational resources, making it difficult to analyze and store the data efficiently.^8,9^

To decrease the size of HSI datasets without losing spectral information, a number of computational techniques have been developed.^9-11^ The simplest method includes spectral binning that involves aggregating adjacent spectral bands that contain similar information. This method has straightforward interpretation and with an additional benefit of removing random noise that is averaged across the selected bin ^12^. However, in this method spectral bands that contain distinct spectral features of different materials are binned together and the resulting broader band represents a mixture of these features. This can complicate the interpretation of spectral signatures, making it increasingly difficult to identify particular materials or detect distinct characteristics.

Other widely used statistical techniques that lead to the compression of hyperspectral data includes Principal Component Analysis (PCA)^13-15^ and Independent Component Analysis (ICA).^14,16^ PCA transforms the original set of variables into a new set of principal components (uncorrelated variables or orthogonal), ordered so that the first few retain most of the variation present in all of the original variables. Despite its effectiveness in data reduction as well as also noise removal, PCA has strong limitation which is interpretation of the data. Unlike the original spectral bands, which correspond to specific wavelengths with known characteristics, the principal components are largely complex features (linear combinations of individual features) that have abstract meaning. In contrast to PCA, ICA results are relatively easy to interpret. The method decomposes a multivariate signal into independent non-Gaussian signals (the process often referred to as the “cocktail party problem,”^17^ where for instance, one needs to isolate individual voices from a mixture of sounds at a party). ICA identifies the most basic independent from each other elements in the dataset that, when combined, reconstruct the original data with fewer components thus achieving the compression. The limitation of ICA lies in the determining the number of independent components to retain without losing critical information. Selecting too few components results in the loss of potentially useful information, and selecting too many introduces noise and redundancies. ICA also relies on the assumption that the spectral signatures of different materials are statistically independent. When this assumption is not supported, such separation into independent components will provide meaningless results.

Deep Learning (DL) algorithms for HSI compression have seen a substantial rise in popularity over the past few years.^18^ Compression via machine learning where a neural network is designed to learn an efficient representation (a.k.a encoding) for a set of data that require dimensionality reduction has been established.^18^ Several spectral signals compressor network based on deep convolutional autoencoder (SSCNet) have been suggested for hyperspectral remote sensing applications.^19^ Such approaches however require a large number of hyperspectral datasets during training to optimize parameters and reach a desirable compression performance. For example recently a large-scale hyperspectral benchmark dataset made up of thousands of non-overlapping image patches has been presented,^20^ however, this and other DL approaches are highly tailored to specific groups of hyperspectral sensors, limiting their generalizability across different sensor types.

Wavelet transform (WT) has recently been applied for hyperspectral data compression primarily in the spatial coordinates.^21^ The use of wavelets in HSI was motivated by the success of JPEG compression, which relies on spatial WT as its core algorithm.^22^ While most WT techniques apply spatial wavelet transforms to reduce the number of pixels maintaining image quality, we envisioned that extending this approach to the spectral domain only will decrease the datasize while preserving essential spatial details. This will be particularly critical in applications such as medical imaging, where spatial information plays a vital role in identifying small lesions and abnormalities. In addition to compression, we have explored how different WT can filter noise, enhance image quality, and improve contrast relative to the original image. We also examined whether the WT approach offers advantages over a benchmark HSI compression method such spectral binning.

## 2. MATERIALS AND METHODS / EXPERIMENTAL

### 2.1. Hyperspectral imaging dataset

The HSI dataset was collected with the HSI pushbroom data system in the shortwave infrared (SWIR) spectral range. The SWIR range, typically spanning wavelengths from 900 nm to 2500 nm, was chosen for this study due to its unique advantages in HSI, particularly for biomedical applications. In the context of biomedical imaging, SWIR offers deeper tissue penetration compared to visible or near-infrared (NIR) wavelengths, enabling the detection of subsurface features such as vascular structures or lesions with high contrast.^23^ SWIR photons are less affected by scattering than shorter wavelengths, resulting in clearer, more detailed imaging of biological tissues. The SWIR range also exhibits strong sensitivity to water, protein and lipids absorption, making it suitable for distinguishing between healthy and diseased tissues based on the spectral signatures of these tissue components^24^ In this study, these characteristics were particularly beneficial for capturing the spectral and spatial differences between lesion sites and healthy skin. From a technical standpoint, SWIR hyperspectral spectra are characterized by relatively broad and smooth spectral features,^25^ which are well-suited for wavelet-based compression methods. Finally, the choice of this range was further supported by the availability of high-performance InGaAs sensors and corresponding optics with adequate sensitivity between 900 and 1650 nm.

The set of hyperspectral images were obtained from an arm of a 50-year-old Caucasian male exposed to poison ivy. This dataset is described in our previous publication.^26^ The original unprocessed dataset has a dimension of 484 x 638 x 510 (y-axis, x-axis, z-spectral axis) featuring 510 spectral bands from 881 nm to 1712 nm.

### 2.2. Image processing

Basic visualization and data processing such as extracting spectra form a single point of from selected ROI, and spectral binning were performed using a built-in-house software IDCubePro^TM^ (licensed from Washington University in St. Louis to HSpeQ LLC, St. Louis, MO). All functions were coded in MATLAB. The workflow of the approach is shown in **Figure 1** and **Figure S1**. The schematics of the scale matching process is shown in **Figure S2**.

**Figure 1.**
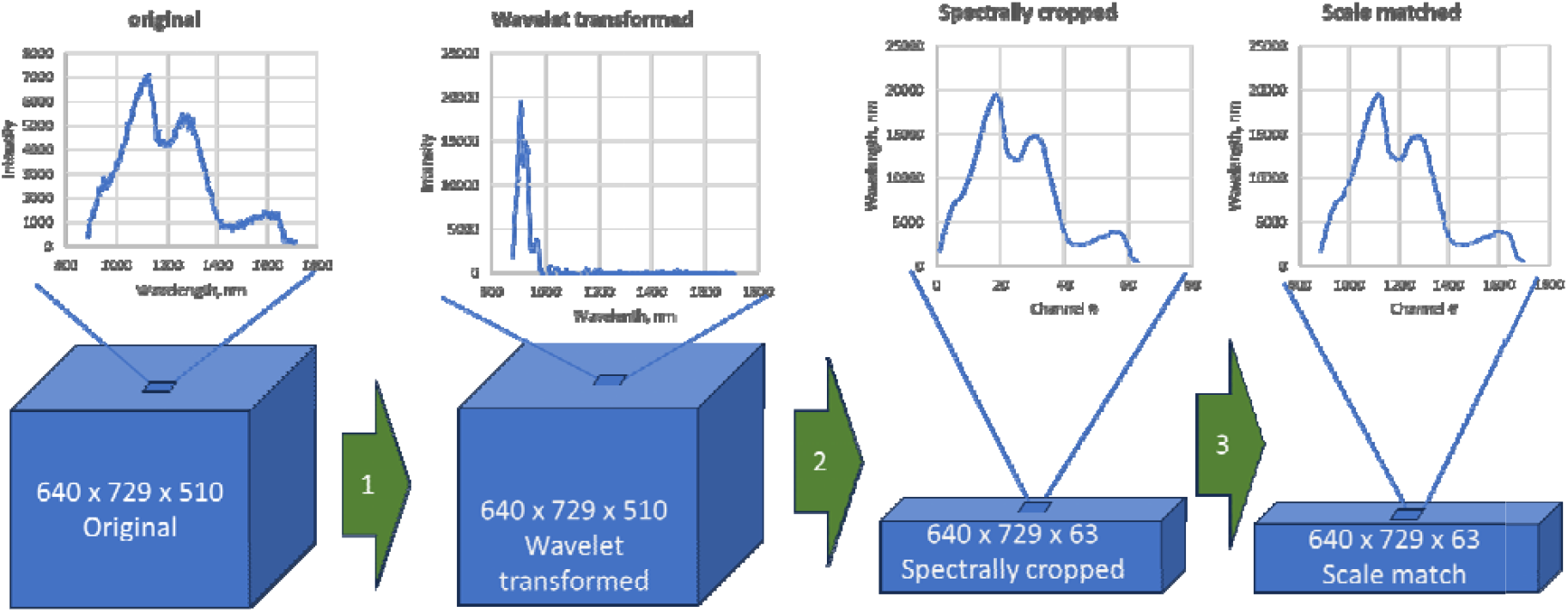
Workflow of the WT compression of a hyperspectral image. The processed invloved three major steps: 1) wavelet transormation on spectral dimension, 2) spectral cropping to remove bands with low intensities, 3) scale matching to assign wavelenghts to the compressed dataset. The axes represent two spatial (x, y) and spectral (z) coordinates

### 2.3. Selection of Daubechies Wavelets and Decomposition Levels

Daubechies (*Db*) wavelets^27^ were selected from other types of wavelets for their compact support and orthogonality, which ensure efficient representation of localized spectral features. The choice of *Db* order (*DbN*) (**Figure S3**) and decomposition levels were guided by their ability to effectively balance data compression and noise reduction.

To identify the optimal wavelet order, a range of wavelet types (*Db1* to *Db10*) was evaluated on their ability to retain the spectral similarity between the original and compressed data. Decomposition levels were selected based on the desired compression ratio and the need to retain critical spectral information. Each decomposition level corresponds to a finer separation of frequency components, with higher levels progressively compressing the data by removing noise. In this study, decomposition levels ranging from 1 to 5 were tested, achieving compression ratios of 8x to 32x. Beyond level 5, then number of bands after compression became small (less than 10), artifacts and spectral distortions became prominent.

### 2.4. Spectral compression with wavelet transform (WT)

The WT compression process was performed in three steps. In the first step, the WT was applied to the spectral dimension of the hyperspectral image (HSI) dataset on a pixel-by-pixel basis. Different types of *Db* wavelets from *Db1* to *Db10*, each with varying decomposition levels, were employed for this process. Following the transformation, in step 2, the dataset was spectrally cropped by removing the bands with close to zero coefficient intensity values. In this step, wavelength values were replaced with the channel numbers (channels #1, #2, #3 etc.). In a third step, a scale-matching algorithm was applied to convert the channel numbers back to their corresponding wavelengths, resulting in a compressed dataset that preserved the correct spectral scale.

### 2.5. Channel-to-Wavelength Matching

Wavelets conversion replaces a wavelength scale to the channel number scale (#1, #2, #3, etc). To restore the wavelength scale, we developed a function that establishes a mapping between spectral channels from a compressed dataset and their corresponding wavelengths from an original dataset. Data from the original file, including wavelengths and intensities, were first smoothed with a moving average filter using three adjusted bands. The largest intensity peak in both datasets is identified, along with the first and last data points. These points were used to perform a linear fit using these three anchor points using *polyfit* function in MATLAB. Using this fit, wavelengths were mapped to channels in the compressed dataset. Intensities from the compressed dataset were interpolated to align with the wavelengths of the original dataset for comparison. Both spectra were normalized, and their similarity is quantified using correlation and root-mean-square error (RMSE). The example of this approach is shown in **Figure S2**. Visualizations of the original, compressed, and mapped spectra, along with their normalized comparisons, are generated using MATLAB’s UI capabilities.

### 2.6. Spectral binning

Spectral binning was performed by combining and averaging the intensities of adjacent spectral bands in the entire dataset to reduce the number of bands and achieve the compression level equal to 8x, 16x and 32x. (see Supplemental Data, **Figure S7**).

### 2.7. Spectral Similarity

The spectral similarity between the regions of interest (ROIs) of the original dataset vs WT compressed dataset was evaluated using the Pearson correlation coefficient. A value close to 1 indicates a strong positive correlation, meaning the spectral profiles are highly similar, while a value close to 0 indicates no correlation.

### 2.9. Signal-to-noise ratio (SNR) measurement

SNR was performed by selecting a “signal” ROI, typically in the middle of the lesion and a “noise” ROI from the part of the image not covered by the sample (where arm is resting). The algorithm calculated the mean signal and variance of the noise by flattening the noise region into a one-dimensional array and computing its variance. The SNR was then calculated for each spectral band using the ratio of the square of the mean signal to the noise variance. The result was then converted to decibels (dB) according to **Eq. 1**:

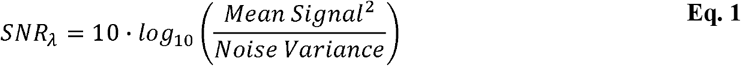

where, *Mean Signal*, is the average value of the signal region for the spectral band λ, *Noise Variance*, is the variance of the noise region for the same spectral band *b*.

### 2.10. Image quality metric

Image quality algorithm calculates the no-reference perceptual image quality score using Blind/Referenceless Image Spatial Quality Evaluator (BRISQUE) method implemented in MATLAB.^28^ The quality score was calculated for each wavelength and presented in a plot Score vs. Wavelength. The range of the quality Score is from 0 to 100, with a smaller score indicating better perceptual quality of the image.

### 2.11. Contrast measurement

Contrast quantification algorithm between two ROIs was based on our published approach^29^ that uses Bhattacharyya distance *D*_*b*_ ^26, 30^ as a metric of contrast. The method is based on the assumption that each ROI is relatively uniform and represented by a single mode histogram representing pixel intensities. ROIs corresponding to the signal and background area were selected from the lesion and the nearby healthy skin. Both ROIs were selected from visually homogeneous areas. The output was a spectrum that represents *D*_*b*_ vs. wavelength. The range for *D*_*b*_ is from 0 (no contrast) to infinity (ideal contrast). Based on our previous experience, areas with low *D*_*b*_ < 0.4 - 0.5 suggest a low contrast with weak difference between the ROIs.^29^ Areas with *D*_*b*_ >3 would indicate a high contrast.

### 2.12. Shannon entropy mapping

To perform entropy-based wavelet compression of hyperspectral images, we developed a MATLAB function (see Code and Data Availability) that processes hyperspectral datacubes by classifying spectral entropy known as Shannon Entropy^31^ and applying multi wavelet decomposition. The Shannon entropy (H) was calculated according to formula **(Eq. 2**.)

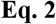

where is the probability of the *i*^*t*h^ spectral band in a given spectrum (normalized intensity values, the normalization step ensures that the sum of all intensity values in the spectrum equals 1, making it a probability distribution), *N* is the total number of spectral bands,

The hyperspectral data was reshaped into a 2D matrix of pixels × bands, and Shannon entropy was computed for each spectrum after normalization. A predefined entropy threshold (=8.7) classified spectra into high-entropy or low-entropy categories, which determined the order of wavelet transform applied.

## 3. RESULTS

To analyze the effects of the WT on data compression, we used the HSI dataset of a human arm affected by poison ivy imaged using a benchtop hyperspectral SWI imager ^26^ To identify the best compression, we applied a variety of *Db* wavelets (shown in **Figure S1**) to the spectral dimension of the HSI dataset on a pixel-by-pixel basis and analyzed the new compressed datasets using established metrics such as SNR, image quality, and contrast between the lesion and the healthy tissue. These metrics were also applied to the spectrally binned dataset, that we used as a standard benchmark for HSI data compression, to assess any advantages of WT over this benchmark method.

The schematic of the three-step compression process is shown in **Figure 1** and in more details in **Figure S1**. Various *Db* wavelets with different decomposition levels were used to identify the optimal compression without sacrificing significant information.

In the first step, the spectra from each pixel were decomposed into a set of wavelet coefficients using WT, which separated each spectrum into different frequency components: low-frequency (longer wavelengths) and high-frequency (shorter wavelength). The resulting shorter wavelengths represent the majority of the image’s main features and have larger coefficients intensities, while longer wavelengths components (representing noise) have the coefficients intensities close to zero. Such arrangement generates a spectrum that appears to be “smooshed” toward the lower wavelengths (see **Figure 1**, Wavelet transformed, and **Figure S2**).

In the second step, the resulting spectra were cropped to remove wavelengths with coefficient intensities close to zero. This cropping was performed with several iterations to maximize Pearson correlation with the original spectrum. For this we selected representative pixels from the lesion from the same location (single pixel or ROI) in both the original and WT processed sets. To avoid noisy peaks, we reduced noise in the original dataset using a moving average filter (sliding window) that averaged three adjacent bands.

In the last step, the channels numbers were matched with the corresponding wavelengths using three reference points. The channel #1 was assigned to the second reference point, the reference point #2 corresponded to the largest peak in both the original denoised spectrum (after the sliding average procedure) and the post cropped spectrum, the last channel in the spectrum corresponded to the third channel. The linear equation from the *polyfit* algorithm (see Channel-to-Wavelength Matching in the Methods) was then applied to the entire range of channels, quantifying the goodness of fit with the Pearson correlation coefficient and replacing channel numbers with the corresponding wavelengths. The goodness of fit between the original and compressed spectra were served as an optimization handle for matching. The algorithm maximized this correlation by adjusting the range of channels by selectively removing channels from both ends of the compressed spectrum. The example of the resulting spectrum using wavelet *Db3* with a decomposition level 3 (*Db3_3*) is shown in **Figure 2** and the entire process is shown in **Figure S2**. Such process results in 8x time smaller dataset with no apparent changes in the spectra shape compared to the original spectrum.

**Figure 2.**
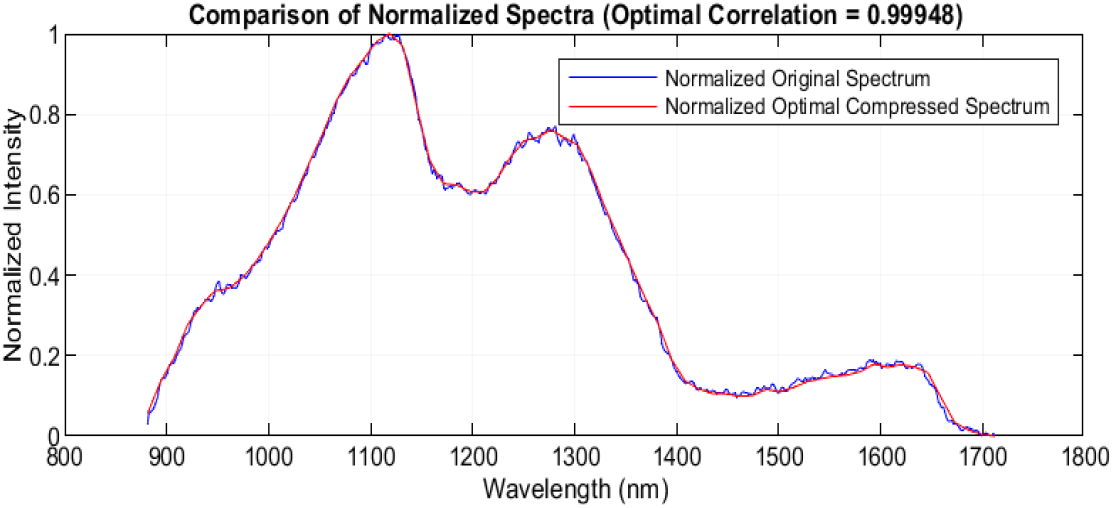
Example of the matching scale for a compressed spectrum using automatic channel-to-matching algorithm. The compressed spectrum with matched wavelengths produced a strong match the original spectrum with Pearson correlation coefficient *q*= 0.99948. Shown as an example is a *Db3* type wavelet with a decomposition level 3 (*Db3_3*) that provides 8x compression.

### 3.1. Daubechies (Db) wavelets

As shown in **Figure S3**, *Db* wavelets differ in complexity which influences their ability to capture detail and smoothness in data. We used *Db* family of wavelets due to their compact support, which makes them highly effective for representing localized spectral features. Compact support in wavelets refers to a property of a function being nonzero only within a finite interval, ensuring its influence is confined to a specific, relatively small region.^32^ In the hyperspectral context, this property means the wavelet transforms a limited number of adjacent spectral bands at any given point, while ignoring bands outside this range. For different *Db* specified with the order *N*, (*DbN*), the number of adjacent bands is approximately proportional to *2N−1*, where *N* is the wavelet order. For example, *Db1* affects 2 adjacent bands, *Db2* affects 4 adjacent bands, *Db3* affects 6 adjacent bands and so on. Lower-order wavelets (e.g., *Db1, Db2*) are shorter and better at capturing sharp, local details, making them suitable for applications that require fine resolution. In contrast, higher-order wavelets (e.g., *Db5, Db6*) are longer and are better suited at capturing broader features with greater smoothness. This compact localization allows *Db* wavelets to focus on essential spectral information, such as peaks and transitions, while discarding redundant or less significant components. A general trend for the given SWIR HSI dataset with relatively broad bands and smooth transitions, is that the optimal order apparently lies between 2 and 4 with the decrease of the spectral preservation at higher order as shown in **Figure S4**.

### 3.2. Decomposition levels in spectral compression

The choice of the wavelet is important for high quality WT; however, it is the decomposition level that directly impacts data compression and the balance between detail retention and noise reduction. The decomposition level defines how many times the WT is applied. In the context of spectral compression using WT, the decomposition level refers to the number of times the spectral data is iteratively broken down into lower- and higher-frequency components. Each level progressively compresses the spectral data by reducing its dimensionality while trying to preserve essential spectral information as shown in **Figure S5**.

At lower decomposition levels (e.g., Level 1 or 2 as in *Db3_1* and *Db3_2*), the WT captures more of the high-frequency components, retaining fine spectral details such as sharp peaks and narrow bands. These levels are useful for maintaining high spectral resolution, ensuring that small but significant variations in the spectrum remain intact. However, compression at lower levels results in less data reduction, which may still leave some noise or redundant information in the compressed spectra. At higher decomposition levels, the WT compresses the spectrum more aggressively by focusing on broader, low-frequency components. This helps to filter out high-frequency noise and reduces the amount of data significantly, but also lead to the loss of finer spectral details.

As shown in **Figure 3**, all *Db* wavelets from *Db1* to *Db10* exhibit a high level of spectral similarity with the original spectrum at all orders, with correlations greater than 0.995 for decomposition levels up to 4 corresponding to 16x compression. Starting from level 5 (compression 32x), the effect of higher decomposition levels becomes more pronounced, and differences between wavelet orders become more apparent. Notably, at this level, *Db3_5* shows the highest correlation compared to others *DbN_5*. Importantly, all *Db* wavelets demonstrate significantly higher correlations with the original spectrum compared to spectral binning, as indicated by the dotted line in **Figure 3a**. However, at this high level the compressed spectrum started showing some artifacts such as around 1600 nm (**Figure 3b)**.

**Figure 3.**
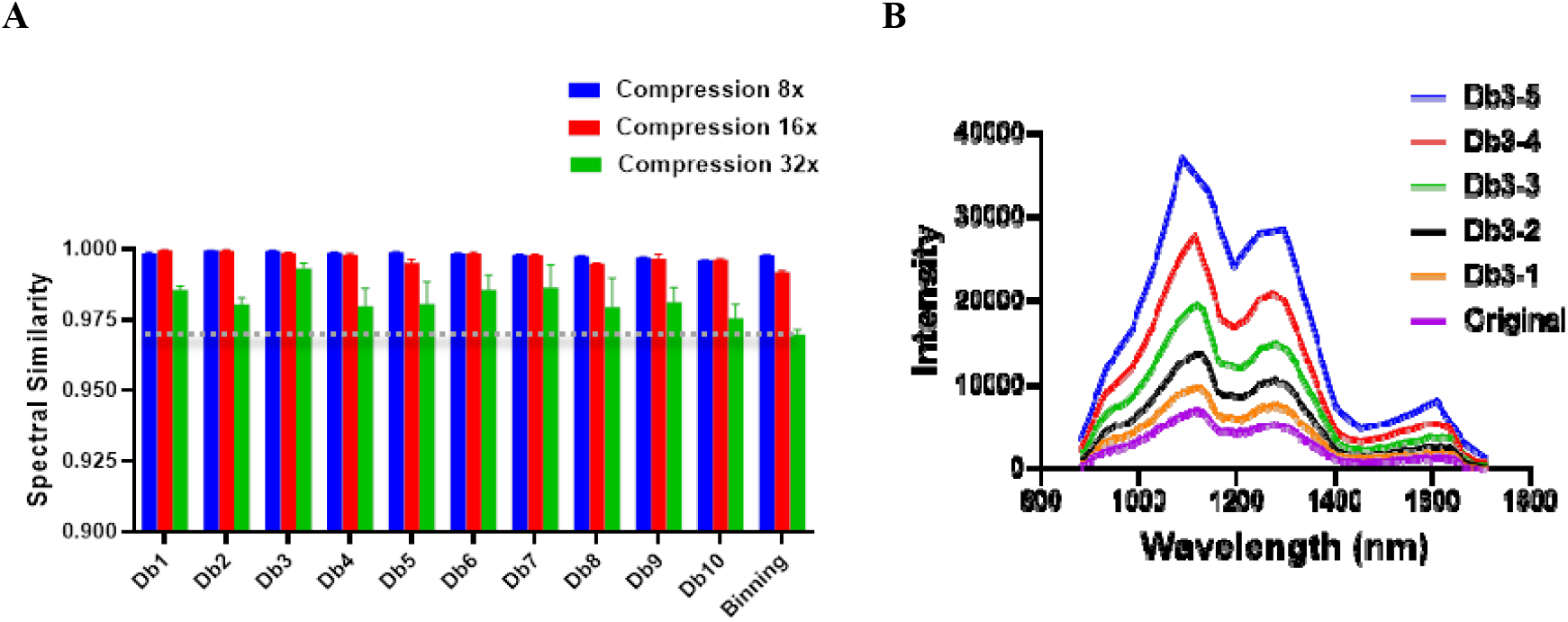
Effect of *Db* types with different levels of decomposition. **A:** Spectral similarity of *Db* wavelets within the same ROI at three decomposition levels: *DbN_3, DbN_4, and DbN_5* (*N* denotes wavelet order), corresponding to compression levels 8x, 16x, and 32x respectively, compared with spectral binning at matching compression levels. Error bars reflect different location in the image and correspond to n = 3 for each datapoints. Dotted line shows the level of Spectral Binning at 32x compression. **B**: spectra of *Db3_M* at different levels of decomposition.

### 3.3. Identifying the effective wavelets using spatial quantitative metrics: SNR, quality and spatial contrast

The produced compressed datasets were then analyzed by three metrics to evaluate the potential loss of the data quality un the spatial domain. We calculated the SNR using the lesion as the ‘signal’ and the stage reflection as the ‘noise’. We assessed the quality of the entire image for each wavelength using BRISQUE algorithm. We measure the contrast between the lesion and the nearby healthy tissue using Bhattacharyya distance. To establish consistent measurements, the areas were selected from the same coordinates.

#### SNR

SNR was performed by selecting an area of the representative signal, typically in the middle of the lesion and in the representative noise area in the background as shown in **Figure 4a**. WT improved the SNR as shown in **Figure 4b**. This improvement is largely due to the transform’s ability to separate noise and signal across different frequency bands similar to the Fourier analysis. Higher SNR values generally correspond to better data quality. Low SNR below 950 nm and above 1650 nm point to the limitation of the sensor InGaAs sensitivity sensor that was used in the SWIR camera. WT improved this sensitivity.

**Figure 4.**
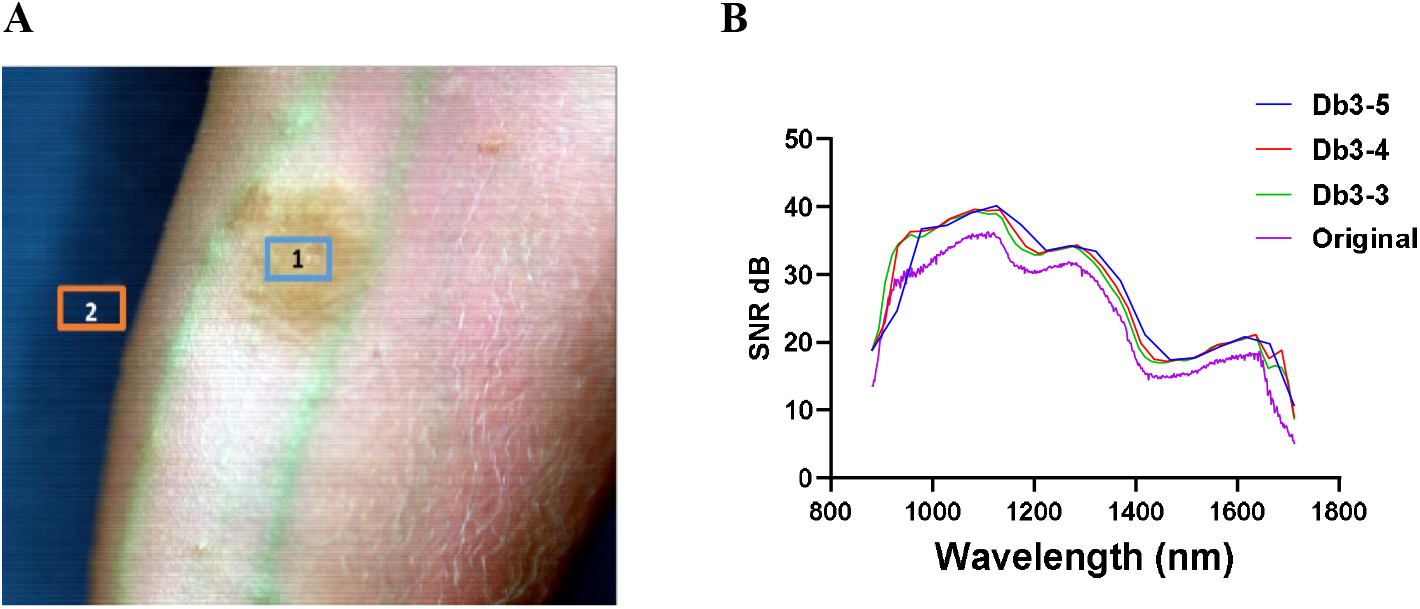
Signal-to-noise ratio (SNR) depends on the level of decomposition. **A:** The original pseudo-RGB image shows regions of interest (ROIs) for the signal (lesion, denoted as Box 1) and the noise (background outside the hand, denoted as Box 2). **B:** The calculated SNR for each wavelength is presented as a spectral plot. Note that the SNR is lowest around 1550 nm, where water absorption is highest, and beyond 1650 nm, where the detector sensitivity decreases.

Optical signals such as seen in biological systems tend to vary more smoothly over time, concentrating their energy in lower frequencies. In contrast, noise, particularly white noise, is characterized as being random and uncorrelated across time. This randomness leads to rapid and frequent fluctuations in the signal. White noise is often present in higher-frequency components and can be attenuated through thresholding. WT allows for noise reduction while retaining localized features of the signal. This selective filtering effectively reduces the noise floor and enhances the clarity of important signal components, leading to an overall improvement in SNR as seen in **Figure 4b**. Interestingly, the SNR values after WT were slightly higher that the SNR after spectral binning (**Figure S7a**) although the difference was marginal.

#### Image quality

BRISQUE (Blind/Referenceless Image Spatial Quality Evaluator) method implemented in MATLAB, calculates the perceptual quality score of the image.^28^ This method is performed by a model trained on an image database that contains images with known distortion such as compression artifacts, blurring, and noise, as well as original versions of the distorted images. The algorithm captures how the image deviates from expected quality. Although BRISQUE was initially developed for natural scenes, we rationalized that it can be effectively applied to assess the quality of any type of image, including monochromatic slices from hyperspectral datasets. Herein, we applied BRISQUE to assess images at individual spectral bands across the entire dataset.

The results shown in **Error! Reference source not found.6** demonstrate the advantages of WT in improving the quality of hyperspectral data and minimizing deviations within the dataset. The original dataset exhibits a relatively large variation in quality between the frames, with the image quality significantly decreasing (higher BRISQUE score) in regions where the camera’s spectral sensitivity is low—below 950 nm and above 1650 nm. Interestingly, the image quality of the original set is somewhat better around 1450 nm, which is likely due to higher absorption by water from biological tissue and, therefore, less scattering that causes blurriness. Application of WT to the original dataset resulted in more uniform quality across the entire spectral range, without significant loss of image quality at most wavelengths and with noticeable improvements at the edges of the spectrum. The results were in general similar to binning (**Figure S7b**), except at longer wavelengths above 1600 nm, where WT showed better score.

#### Spatial contrast

The Bhattacharyya distance was used for the assessment of the spatial contrast. This approach described in our previous publications^29, 30^ measures the contrast between two regions of interest (ROIs). This metric quantifies the degree of overlap between the probability distributions of pixel intensities (or other relevant features within the two ROIs). A lower Bhattacharyya distance indicates a higher overlap (i.e., lower contrast), while a higher distance suggests greater separability between the ROIs, reflecting better contrast. The results shown in **Figure 5** demonstrate an increase of the contrast between the lesion and healthy tissue across almost the entire spectral range for the level of decomposition 3 and 4 (*Db3_3*, and *Db3_4*). At the level 5 (*Db3_5*) the contrast largely returns to the original level. In contrast, spectral binning with the same level of compression largely retains the same contrast level as the original from the 8x and 16x compression, although increased in 32x (*Db3_5*) compression (**Figure S7c**). This data suggests that even at this high level of WT compression, the contrast between objects are not negatively affected.

**Figure 5.**
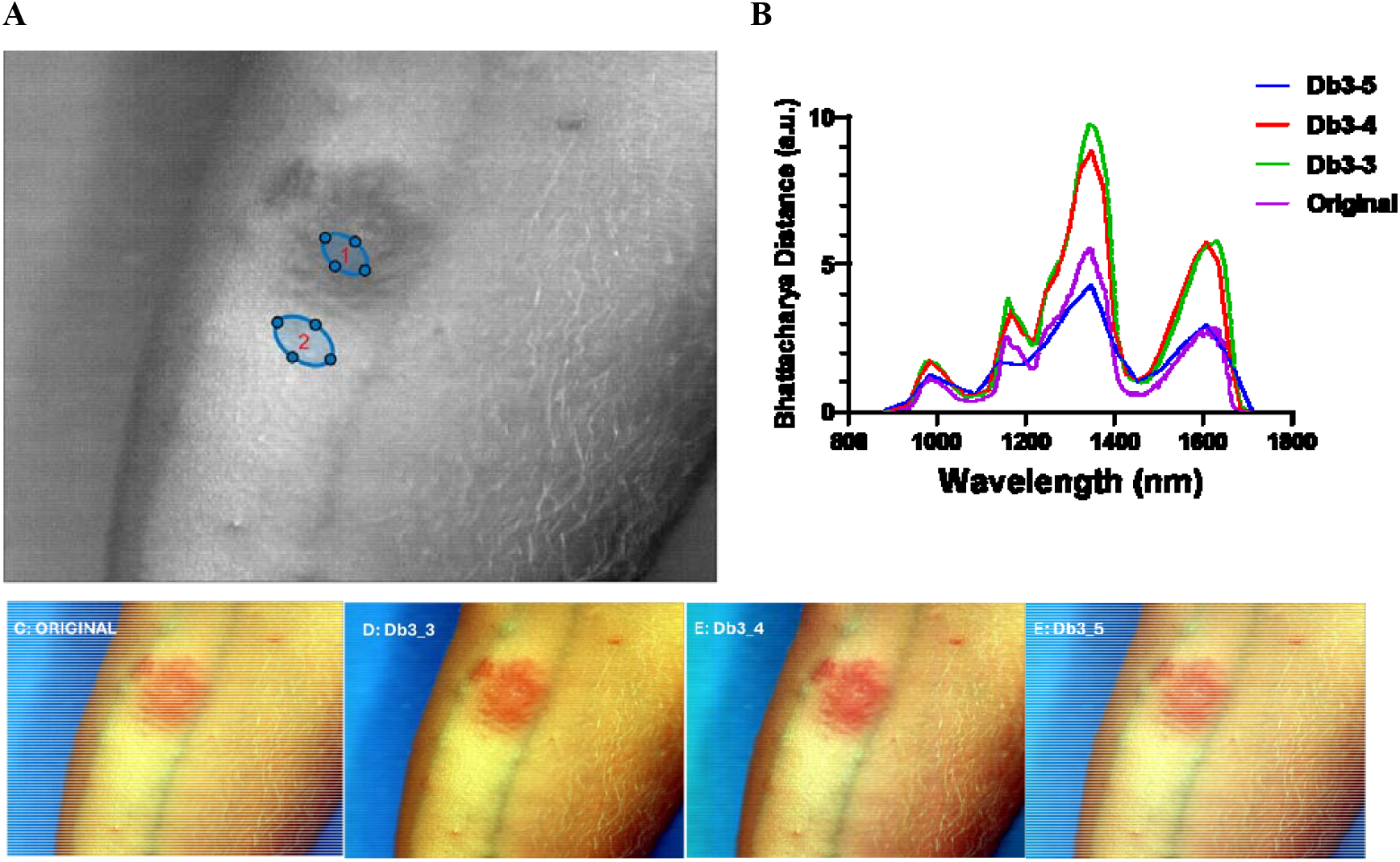
Contrast measurement. **A:** selection the region of interest (lesion, area 1) and healthy skin (area 2). **B**: calculating Bhattacharyya distance (*D*_*b*_) between two areas across all wavelengths in the datacube. *D*_*b*_ values above 3 can be considered as highly contrasted, while values below 0.5 typically show no contrast. **C-F**: pseudo RGB images of the original and after WT processing. The spatial resolutions for the pseudo RGB images are almost identical (also see **Video S1** to visualize small changes between the original and compressed image) for all cases while the contrast is slightly changes for ROIs 1 vs 2.

### 3.4. Mixed wavelet approach

Hyperspectral datasets present heterogeneous spectral information, with varying degrees of complexity across different regions. In the studied dataset, pixels correspond to distinct classes, including injured tissue, normal skin, and the background, which represents the stage. Each of these regions exhibits unique spectral characteristics and different spectral complexity due to the presence of multiple biochemical and material components.

Given this variability, a multi WT approach to spectral processing can be beneficial. Instead of applying a single wavelet transform uniformly across the dataset, we pre-classified pixels based on their spectral complexity. We envision that Shannon entropy,^31^ which quantifies the level of spectrum complexity, allows differentiation between regions with different spectral signatures Indeed, as shown in **Figure 6**, the entropy can distinguish between the background and the tissue, although this difference seems relatively small dynamic range. Nevertheless, the threshold of 8.70 can effectively separate the two types of data, where the low-entropy regions belong to the tissue, and the higher entropy belong to the background. This is contradictory to the expectation that higher-entropy regions reflect higher spectrum variability, such as tissue, with multiple spectral components. The comparison between the single WT (*Db3_3*) and multi WT approach when *Db1_3* was applied to the high entropy regions and *Db3* for the low entropy areas showed an improvement in SNR at wavelengths between 1550 and 1650 nm, although at the expense of lower SNR between 940 nm and 1140 nm. The opposite approach, when *Db3_3* for high entropy areas and *Db1_3* for low entropy areas showed an opposite trend: the increase SNR between 940 and 1140 nm and decrease at wavelengths longer than 1650 nm

**Figure 6.**
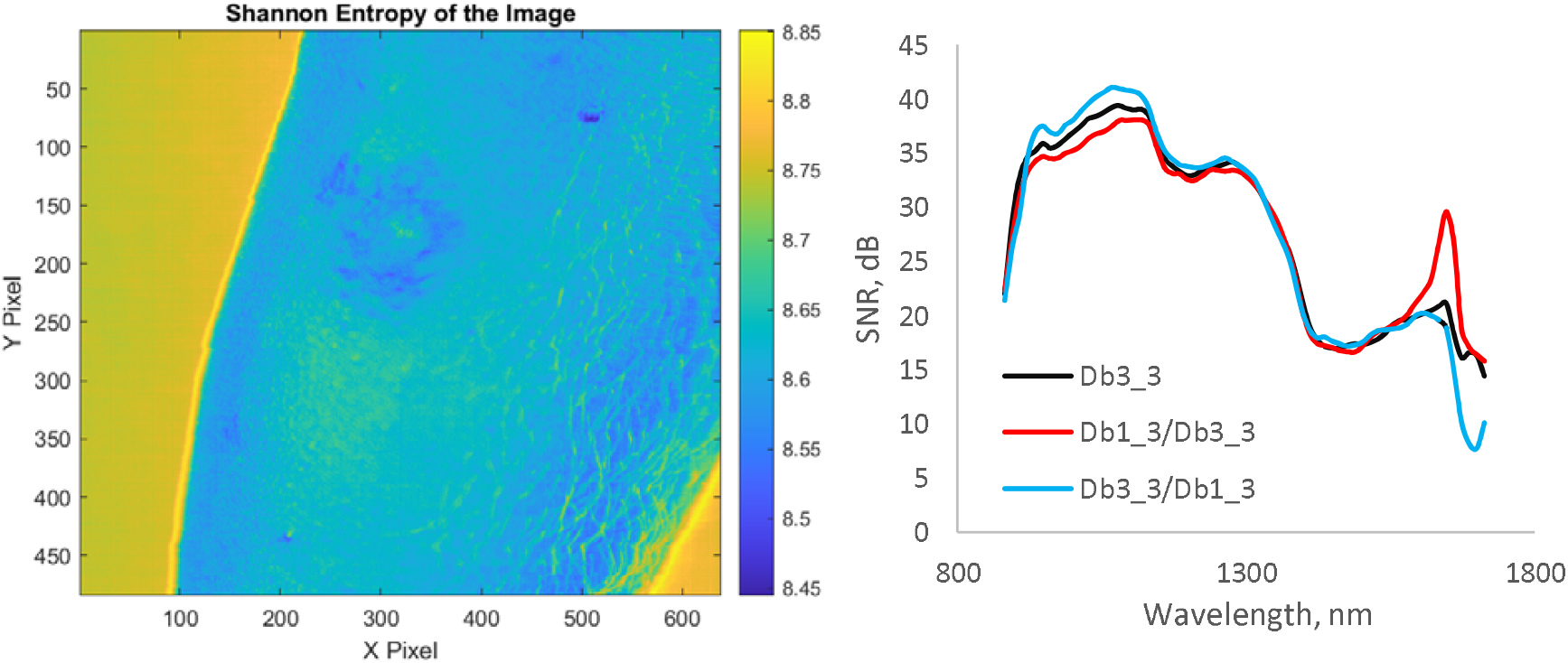
Pre-evaluation of pixels for wavelet analysis based on entropy of spectral. **A:** Shannon entropy map of the hyperspectral dataset, illustrating spectral complexity across different regions. Higher entropy values correspond to the background, and lower entropy values represent the arm. **B**: Effect of a mixed wavelet approach on SNR between the skin and the background. *Db3_3* corresponds to the WT with order 3, and the level of decomposition equal 3. *Db1_3/Db3_3* corresponds to the WT where the *Db1_3* was applied to the yellow part of the map above threshold 8.7, while *Db3_3* was applied to the blue part of the map (<8.7). *Db3_3/Db1*_3 corresponds to the WT where *Db3_3* was applied to the yellow part of the map above threshold 8.7, while *Db1_3* was applied to the blue part of the map (<8.7).

## 4. DISCUSSION

WT are often used in various aspects of medical imaging, including noise filtering, feature extraction, and image compression. WT filtering algorithms have been effective in noise reduction for ultrasound images,^33^ feature extraction in MRI and CT scans,^34, 35^ classification of medical images.^36^ and for image reconstruction in tomographic imaging.^37^

In this work we applied WT for compression of hyperspectral data. Our results showed that wavelet compression offers a promising strategy for compressing such data obtained from the bench type imagers. Medium orders of wavelet, such as *Db3* with decomposition levels between 3 to 5, effectively captured the essential, image carrying low-frequency information while reducing data size and filtering noise. This approach retains the critical spectral features despite the significant decrease of the dataset size (up to 32 times). The following discussion compares the WT compression compared to other compression methods and explains the role of various factors in achieving efficient compression without the loss of significant data.

### 4.1. Wavelet order defines separation of signal from noise while the level of decomposition is critical for compression

Wavelets come in many families, each suited to different types of signal analysis. *Db* family wavelets (*Db* family), named after Ingrid Daubechies^27^ (**Figure S3**), are among the most widely used. As mentioned above, low-order wavelets (such as *Db1* or *Db2*) have shorter support, allowing them to better capture sharp transitions and fine details. In contrast, higher-order wavelets effectively capture smooth parts of a signal, making them suitable for representing gradual trends.

In HSI of 3D objects (i.e. small animals, humans’ body parts, etc), spectral data have complex baselines due to factors like uneven illumination, sample curvature, surface roughness, chromatic aberration and varying tissue scattering across wavelengths. While baseline minimization techniques—such as subtracting reflectance from reference materials (e.g., using a Spectralon strip) or accounting for dark counts—can partially reduce these effects, challenges remain. Optimal *Db* effectively address these issues by filtering out complex baseline patterns without directly including the baseline into the model.

While the order of the *Db* wavelet function is important for efficient separation of the signal from noise or baseline, the level of decomposition *M* denoted as *DbN_M* plays an especially important role in data compression. This parameter defines how many times that wavelet is applied concurrently and hence the compression level appears close to be proportional to the power of 2 (**Eq. 3**).

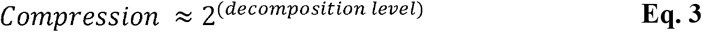

Decomposition in WT is a process that involves breaking spectral data set into different frequency subbands. Higher levels of decomposition lead to better compression ratios. However, the trade-off for using relatively high levels of decomposition for hyperspectral data is that it rapidly decreases the number of non-zero bands. For example, in *DbN_6* the total number of remaining bands in the shown case with the original 510 bands reduced to only 8 bands, that dissipates the advantage of high spectral resolution and therefore classification and identification of objects based on the spectral features.

### 4.2. *Db*-order and decomposition levels are independent parameters

The *Db* order (*N)* and the level of decomposition (*M*) function as independent variables, each contributing uniquely to the analysis process. The choice of *Db* wavelet type specifies the support, which determines the wavelet’s ability to capture several adjacent bands. Conversely, the level of decomposition governs how many times this type of WT is applied. Each level captures progressively coarser trends, with higher levels isolating larger-scale (lower-frequency) features within the data. This layering effect allows the decomposition level to operate independently of the *Db*-type wavelet, allowing to adjust both variables to best suit the specific resolution and scale requirements of their analysis. Screening of *Db* from 1 to 10 showed that *Db3* with an appropriate level of decomposition provided an optimal balance (**Figure 3a**) for the given experimental dataset, resulting in spectra that closely matched the original with maximum correlation.

There are two distinct methods for applying a *DbN_M* WT: i) one-time application of *DbN_M*, ii) *M-*times *applications of DbN*, and iii) *M-times adaptive DbN*. The first, presented in this study *DbN_M*, is a single, M-level decomposition with *DbN*, where each level produces approximation and detail components without selective filtering, resulting in a broad, fixed multi-resolution breakdown of the data. The second method, *M* times *DbN*, involves recursively applying *DbN M* times, with each step decomposing only the low-frequency component from the previous level. This recursive application results in a hierarchy of frequency bands, each representing different layers of detail in the spectrum. This is similar to zooming in on the spectrum, where each level of zoom reveals finer details that were not visible at a higher level. In this method all coefficients are retained, so no low-value components are filtered out. The third approach, *M times adaptive DbN*, applies *DbN* recursively with adaptive band-cutting after each step; low-value coefficients are removed before the next decomposition, focusing only on high-information content. This adaptive method can potentially offer superior noise reduction and compression by dynamically removing low-value data. This *M times adaptive DbN* approach might be the most effective, as it selectively retains high-information content while achieving the highest degree of compression. This approach approach will be investigated in the future.

### 4.3. WT vs Spectral Binning

Since both WT and spectral binning can achieve noise reduction and data compression, it is valuable to compare these two techniques and assess whether the more sophisticated WT-based compression offers advantages over the simpler spectral binning approach. Spectral binning reduces data by averaging neighboring spectral bands and replacing each group with a single average value.

Spectral binning is a one-step procedure, while our WT-based compression algorithm involves three steps: wavelet transformation, spectral cropping (an optimization procedure in our approach), and wavelength matching, all of that slows image processing. While binning is simpler and significantly faster (0.2 seconds for binning vs. 80 seconds for WT on a standard desktop PC). These points raise a key question: does the added complexity of WT justify the effort, or would a straightforward spectral binning approach suffice for compression?

Our results show the clear benefits of WT for data compression compared to spectral binning at least for the given case. For the same compression levels (16x or 32x), WT-compressed spectra show higher Pearson correlation coefficients with the original spectra, better or equal image quality of individual frames, improved SNR and contrast between nearby ROIs. Specifically, our findings indicate that *Db3_4* (compression level 16x) provides ideal compression for SWIR HSI, not only preserving image quality but also enhancing the quality at some wavelengths. *Db3_5*, at a higher compression level of 32x, largely retains essential spectral details. Furthermore, the WT approach offers adaptability by applying wavelet transformation at the individual pixel level based on the spectral complexity, making it highly flexible. In contrast, spectral binning lacks this level of customization and cannot be easily implemented in a similarly adaptive manner.

### 4.3. Generalizability to Other Spectral Ranges and Clinical Implications of HSI compression

The wavelet-based compression approach might have significant potential for application beyond the SWIR range, extending to the ultraviolet-visible (UV-Vis) regions as well as infrared. The *Db* wavelet family’s flexibility and scalability make it adaptable to datasets with varying spectral characteristics. For example, visible and UV HSI, are commonly used in applications such as dermatological diagnostics, endoscopy, intraoperative imaging, including robotics, and fluorescence imaging. Visible hyperspectral datasets typically exhibit higher spatial resolution but narrower spectral ranges, requiring adjustments to compression algorithms to integrate spatial and spectral compression. In contrast IR imaging dataset have low spatial resolution but broad spectral range with sharper peaks. The adaptability of WT to a wide array of spectral ranges applications across different spectral domains makes this tool especially viable.

The large size of hyperspectral datasets presents significant challenges for real-time applications in clinical settings, such as surgical navigation or bedside diagnostics, where rapid data processing and transmission are essential. The wavelet-based compression method enables more than an order of magnitude reducing data size and preserves essential spectral information. This approach ensures that critical diagnostic features, such as spectral signatures of diseased tissues, are maintained. The proposed compression algorithm can be seamlessly integrated with the hardware to avoid time-consuming pre-processing procedures. Furthermore, the method’s ability to enhance SNR and image quality facilitates better visualization of critical structures, making it particularly valuable in detecting early-stage lesions or abnormalities. The efficient storage and rapid processing of the compressed data that retain all pertinent information promises to make this approach integratable with clinical workflows, thus bridging the gap between HSI technology and its practical adoption in healthcare.

## 5. CONCLUSIONS

This study demonstrates that WT offer a highly efficient compression tool for HSI data. By applying *Db* wavelets in the spectral domain, we achieved up to 16x – 32x times compression without sacrificing the critical spectral information necessary for data processing analysis. In comparison to the traditional methods such as spectral binning, WT provides superior contrast, enhanced image quality, and improved SNR - better than the benchmark of compression such as spectral binning. These benefits, combined with acceptable speed and flexibility make WT particularly well-suited for handling the large, complex datasets typical of HSI. The ability to keep the original “wavelength” scale instead of “components” makes WT approach superior to commonly used dimension reduction techniques PCA and ICA that irreversibly loose this important information.

In summary, WT provide a promising approach for advancing the data processing of HSI technology in biomedical field. Future work should explore further optimization of wavelet-based techniques, including the development of custom wavelets for specific imaging applications and extending this approach to a broader range of spectral imaging modalities.

## Supporting information

Supplemental Figures from S1 to S7

Video showing similarity before and after compression

## Data Availability

The dataset and the code are provided in the corresponding depositaries. All other data produced in the present study are available upon reasonable request to the authors.

https://digitalcommonsdata.wustl.edu/preview/b62vrs9x4x?a=ebbf9a22-0909-4b55-a5b7-4e938414c174

## Acknowledgements

The authors thank funds provided by NIH: 1R21NS135646, 5R21CA269099, and 1R01GM141344. The authors thank funds provided by NSF award 1827656. The authors also thank the Optical Spectroscopy Core Facility (NIH original award S10RR031621) at Washington University School of Medicine for computational resources for hyperspectral image analysis.

## Disclosures

Dr. Berezin is the founder of the company HSpeQ LLC, that licensed IDCubePro software from Washington University.

## Code and Data Availability

The code used in this study is available through GitHub at the following URL: https://github.com/MikhailBerezin/Wavelet_hyperspectral

The original and raw data are located in Washington University Depository at the following URL:

https://digitalcommonsdata.wustl.edu/preview/b62vrs9×4×?a=ebbf9a22-0909-4b55-a5b7-4e938414c174

## Biographies

**Mikhail Berezin, PhD** is an Associate Professor of Radiology, Biomedical Engineering and Chemistry at Washington University School of Medicine. He also directs Optical Spectroscopy Core Facility, and he is a Fellow of SPIE. His research interest lies in the development of new optical imaging modalities for peripheral nerve research and associated diseases. His interest also includes the development of contrast agents, fluorescence lifetime imaging, and hyperspectral imaging in SWIR.

**Hridoy Biswas, MS** is a Research Assistant in Radiology at Washington University School of Medicine. His research interests include computational algorithms for hyperspectral imaging, image analysis and machine learning.

**Rui Tang, PhD** is an Instructor in Radiology at Washington University School of Medicine. He received his PhD in Chemistry from Washington University. His research interests include the development of contrast agents and application of optical imaging methods to biomedical applications.

**Shamim Mollah, PhD** is an Assistant Professor at Washington University School of Medicine and a member of the Intelligent Systems in Molecular Biology. She has over 20 years of method development experience in AI technology, NLP, latent space models, graph theory, operation research, and e-discovery. She currently works as Chief Scientific Officer at HSpeQ LLC developing novel directions in hyperspectral imaging.

## Notes

### Funding Statement

NIH funding: 1R21NS135646, 5R21CA269099, and 1R01GM141344.
NSF funding 1827656.

### Author Declarations

The study used published datasets that were submitted in the previous publication: Du, T., Mishra, D. K., Shmuylovich, L., Yu, A., Hurbon, H., Wang, S. T., & Berezin, M. Y. (2020). Hyperspectral imaging and characterization of allergic contact dermatitis in the short‐wave infrared. Journal of biophotonics, 13(9), e202000040. The data are publicly available at the links shown below. The protocol was approved on 12/15/16 by The Washington University in St. Louis Institutional Review Board (IRB ID#201609125).

### Summary of Updates

Added "Mixed wavelet approach: subsection

## References

1. Du, T.; Mishra, D. K.; Shmuylovich, L.; Yu, A.; Hurbon, H.; Wang, S. T.; Berezin, M. Y., Hyperspectral imaging and characterization of allergic contact dermatitis in the short-wave infrared. J Biophotonics 2020, 13 (9), e202000040.

2. Stuart, M. B.; McGonigle, A. J.; Willmott, J. R., Hyperspectral imaging in environmental monitoring: A review of recent developments and technological advances in compact field deployable systems. Sensors 2019, 19 (14), 3071.

3. Singh, P.; Pandey, P. C.; Petropoulos, G. P.; Pavlides, A.; Srivastava, P. K.; Koutsias, N.; Deng, K. A. K.; Bao, Y., Hyperspectral remote sensing in precision agriculture: Present status, challenges, and future trends. In Hyperspectral remote sensing, Elsevier: 2020; pp 121–146.

4. Fox, N.; Parbhakar-Fox, A.; Moltzen, J.; Feig, S.; Goemann, K.; Huntington, J., Applications of hyperspectral mineralogy for geoenvironmental characterisation. Minerals Engineering 2017, 107, 63–77.

5. Mishra, D.; Hurbon, H.; Wang, J.; Wang, S. T.; Du, T.; Wu, Q.; Kim, D.; Basir, S.; Cao, Q.; Zhang, H.; Xu, K.; Yu, A.; Zhang, Y.; Huang, Y.; Garnett, R.; Gerasimchuk-Djordjevic, M.; Berezin, M. Y., IDCube Lite: Free Interactive Discovery Cube software for multi- and hyperspectral applications. J Spectr Imaging 2021, 10.

6. Nalepa, J., Recent Advances in Multi- and Hyperspectral Image Analysis. Sensors (Basel) 2021, 21 (18).

7. Bian, L.; Wang, Z.; Zhang, Y.; Li, L.; Zhang, Y.; Yang, C.; Fang, W.; Zhao, J.; Zhu, C.; Meng, Q.; Peng, X.; Zhang, J., A broadband hyperspectral image sensor with high spatio-temporal resolution. Nature 2024, 635 (8037), 73–81.

8. Firtha, F.; Fekete, A.; Kaszab, T.; Gillay, B.; Nogula-Nagy, M.; Kovács, Z.; Kantor, D. B., Methods for Improving Image Quality and Reducing Data Load of NIR Hyperspectral Images. Sensors (Basel) 2008, 8 (5), 3287–3298.

9. Ghaffari, M.; Lukkien, M. C. J.; Omidikia, N.; Tinnevelt, G. H.; van Eijk, M. C. P.; Podchezertsev, S.; Jansen, J. J., Systematic reduction of hyperspectral images for high-throughput plastic characterization. Sci Rep 2023, 13 (1), 21591.

10. Dua, Y.; Kumar, V.; Singh, R. S., Comprehensive review of hyperspectral image compression algorithms. Optical Engineering 2020, 59 (9), 090902–090902.

11. Motta, G.; Rizzo, F.; Storer, J. A., Hyperspectral data compression. Springer Science & Business Media: 2006.

12. Dell’Endice, F.; Nieke, J.; Koetz, B.; Schaepman, M. E.; Itten, K., Improving radiometry of imaging spectrometers by using programmable spectral regions of interest. ISPRS Journal of Photogrammetry and Remote Sensing 2009, 64 (6), 632–639.

13. Ferrari, C.; Foca, G.; Ulrici, A., Handling large datasets of hyperspectral images: reducing data size without loss of useful information. Anal Chim Acta 2013, 802, 29–39.

14. Mantripragada, K.; Dao, P. D.; He, Y.; Qureshi, F. Z., The effects of spectral dimensionality reduction on hyperspectral pixel classification: A case study. PLoS One 2022, 17 (7), e0269174.

15. Zabalza, J.; Ren, J.; Ren, J.; Liu, Z.; Marshall, S., Structured covariance principal component analysis for real-time onsite feature extraction and dimensionality reduction in hyperspectral imaging. Appl Opt 2014, 53 (20), 4440–9.

16. Boukhechba, K.; Wu, H.; Bazine, R., DCT-Based Preprocessing Approach for ICA in Hyperspectral Data Analysis. Sensors (Basel) 2018, 18 (4).

17. Lee, T.-W.; Ziehe, A.; Orglmeister, R.; Sejnowski, T. In Combining time-delayed decorrelation and ICA: Towards solving the cocktail party problem, Proceedings of the 1998 IEEE International Conference on Acoustics, Speech and Signal Processing, ICASSP’98 (Cat. No. 98CH36181), IEEE: 1998; pp 1249–1252.

18. Afrin, A.; Mamun, M. A. In A Comprehensive Review of Deep Learning Methods for Hyperspectral Image Compression, 2024 3rd International Conference on Advancement in Electrical and Electronic Engineering (ICAEEE), 25-27 April 2024; 2024; pp 1–6.

19. La Grassa, R.; Re, C.; Cremonese, G.; Gallo, I., Hyperspectral data compression using fully convolutional autoencoder. Remote Sensing 2022, 14 (10), 2472.

20. Fuchs, M. H. P.; Demir, B., HySpecNet-11k: A Large-Scale Hyperspectral Dataset for Benchmarking Learning-Based Hyperspectral Image Compression Methods. arXiv preprint 2306.00385 2023.

21. Báscones, D.; González, C.; Mozos, D., Hyperspectral Image Compression Using Vector Quantization, PCA and JPEG2000. Remote Sensing 2018, 10 (6), 907.

22. Usevitch, B. E., A tutorial on modern lossy wavelet image compression: foundations of JPEG 2000. IEEE signal processing magazine 2001, 18 (5), 22–35.

23. Zhang, H.; Salo, D.; Kim, D. M.; Komarov, S.; Tai, Y.-C.; Berezin, M. Y., Penetration depth of photons in biological tissues from hyperspectral imaging in shortwave infrared in transmission and reflection geometries. Journal of biomedical optics 2016, 21 (12), 126006–126006.

24. Cao, Q.; Zhegalova, N. G.; Wang, S. T.; Akers, W. J.; Berezin, M. Y., Multispectral imaging in the extended near-infrared window based on endogenous chromophores. Journal of biomedical optics 2013, 18 (10), 101318–101318.

25. Gruensfelder, H. D.; Shofu, F.; Michie, M. S.; Berezin, M. Y.; Shmuylovich, L.; O’Brien, C. M., Characterization of Biological Absorption Spectra Spanning the Visible to the Short-Wave Infrared. Journal of Visualized Experiments (JoVE) 2025, (215), e67403.

26. Du, T.; Mishra, D. K.; Shmuylovich, L.; Yu, A.; Hurbon, H.; Wang, S. T.; Berezin, M. Y., Hyperspectral imaging and characterization of allergic contact dermatitis in the short_Jwave infrared. Journal of biophotonics 2020, 13 (9), e202000040.

27. Daubechies, I., The wavelet transform, time-frequency localization and signal analysis. IEEE transactions on information theory 1990, 36 (5), 961–1005.

28. Mittal, A.; Moorthy, A. K.; Bovik, A. C., No-reference image quality assessment in the spatial domain. IEEE Transactions on image processing 2012, 21 (12), 4695–4708.

29. Kim, D. M.; Zhang, H.; Zhou, H.; Du, T.; Wu, Q.; Mockler, T. C.; Berezin, M. Y., Highly sensitive image-derived indices of water-stressed plants using hyperspectral imaging in SWIR and histogram analysis. Scientific Reports 2015, 5 (1), 15919.

30. Goudail, F.; Réfrégier, P.; Delyon, G., Bhattacharyya distance as a contrast parameter for statistical processing of noisy optical images. J. Opt. Soc. Am. A 2004, 21 (7), 1231–1240.

31. Shannon, C. E., A mathematical theory of communication. The Bell system technical journal 1948, 27 (3), 379–423.

32. Mallat, S. G., A theory for multiresolution signal decomposition: the wavelet representation. IEEE Transactions on Pattern Analysis and Machine Intelligence 1989, 11 (7), 674–693.

33. Vilimek, D.; Kubicek, J.; Golian, M.; Jaros, R.; Kahankova, R.; Hanzlikova, P.; Barvik, D.; Krestanova, A.; Penhaker, M.; Cerny, M.; Prokop, O.; Buzga, M., Comparative analysis of wavelet transform filtering systems for noise reduction in ultrasound images. PLoS One 2022, 17 (7), e0270745.

34. Laine, A. F., Wavelets in temporal and spatial processing of biomedical images. Annu Rev Biomed Eng 2000, 2, 511–50.

35. Pizurica, A.; Philips, W.; Lemahieu, I.; Acheroy, M., A versatile wavelet domain noise filtration technique for medical imaging. IEEE Trans Med Imaging 2003, 22 (3), 323–31.

36. Baharlouei, Z.; Rabbani, H.; Plonka, G., Wavelet scattering transform application in classification of retinal abnormalities using OCT images. Sci Rep 2023, 13 (1), 19013.

37. Rosenthal, A.; Jetzfellner, T.; Razansky, D.; Ntziachristos, V., Efficient Framework for Model-Based Tomographic Image Reconstruction Using Wavelet Packets. IEEE Transactions on Medical Imaging 2012, 31 (7), 1346–1357.

