## Supplemental Figures from S1 to S7 for "Wavelet-Based Compression Method for Scale-Preserving SWIR Hyperspectral Data"

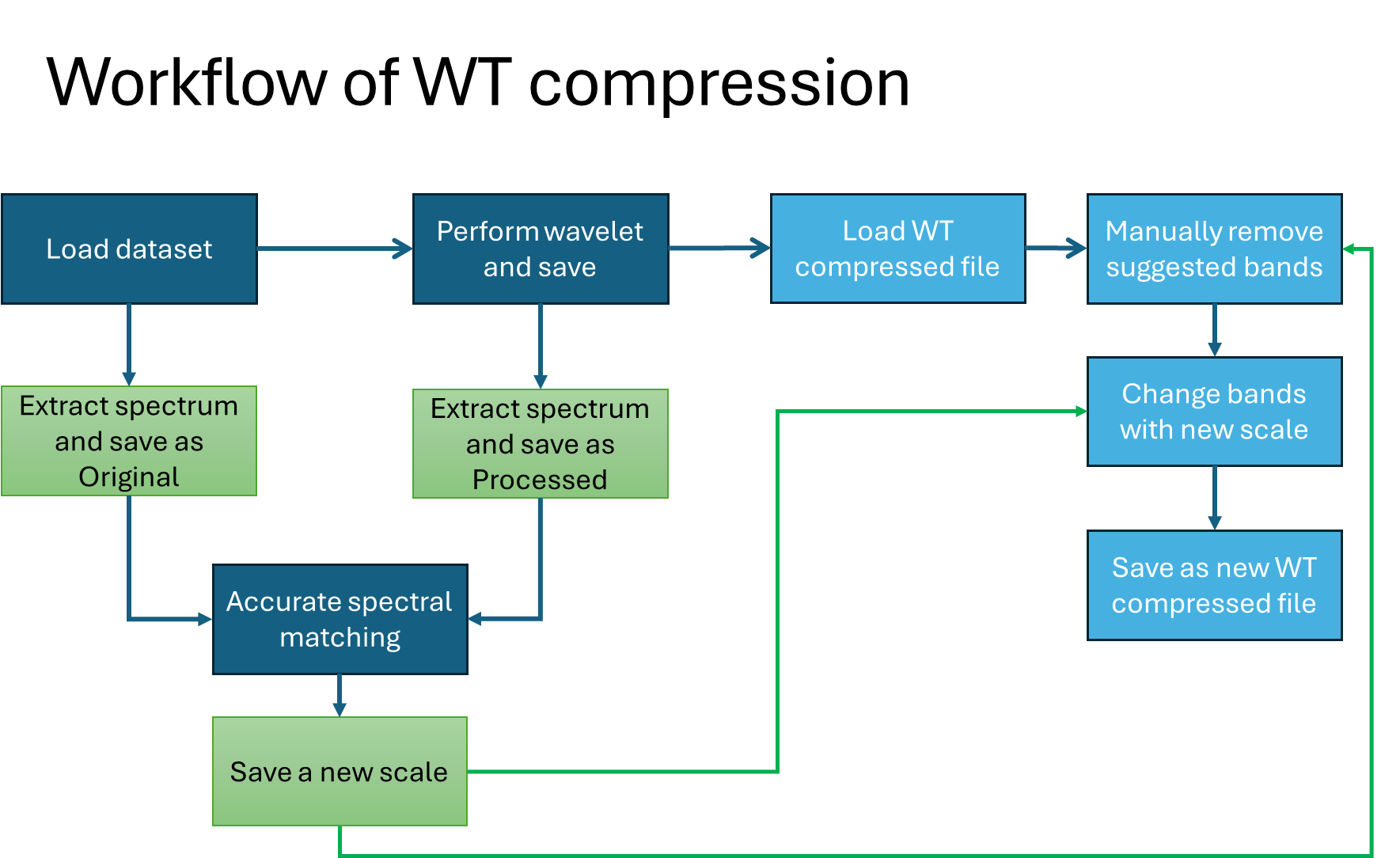

**Figure S1** The workflow of WR compression with spectral matching

| **A**  **B**  **C**  **D**  **E** | 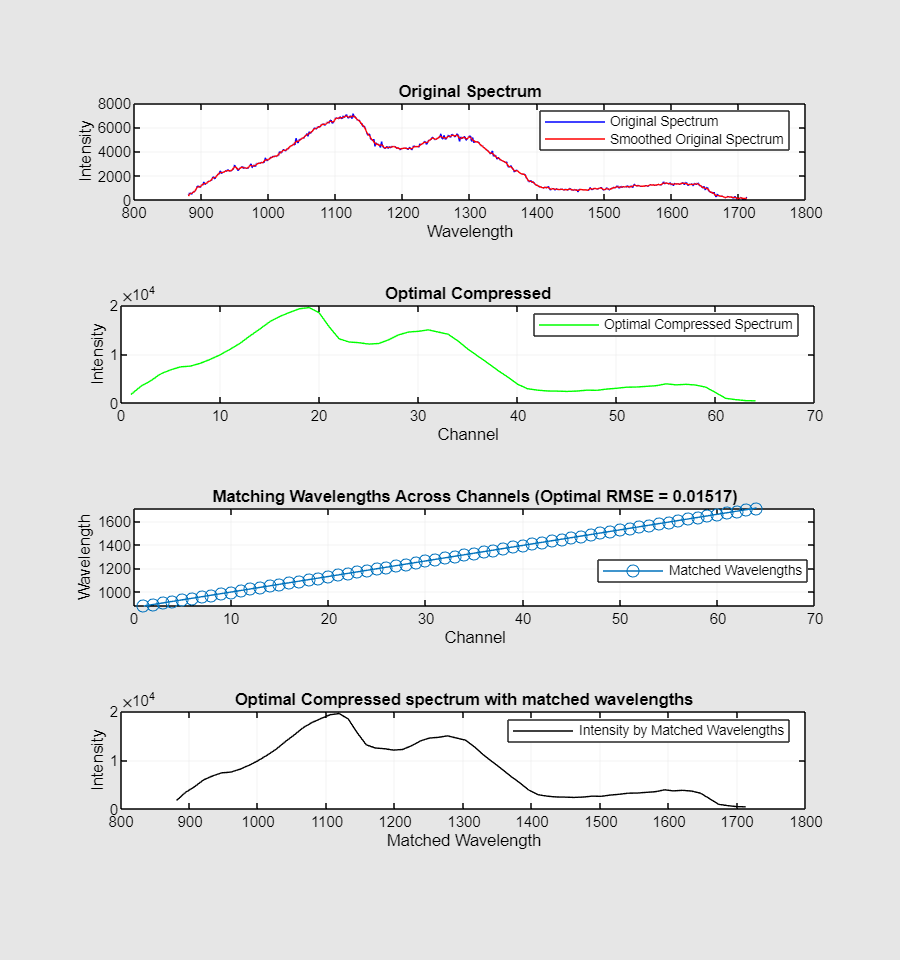  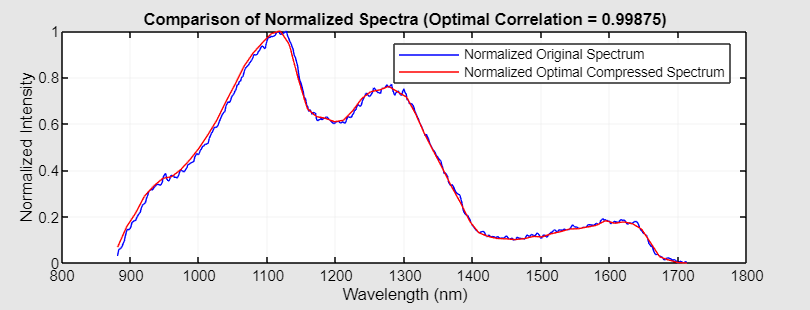 |
| --- | --- |

**Figure S2** **Spectral matching processing. A**: The spectrum from a selected pixel within the dataset is first smoothed with moving average of three adjacent bands to reduce false peaks. **B:** The corresponding spectrum from the wavelet-transformed (WT) compressed dataset at the same spatial position is retrieved. **C:** Wavelength matching is performed using root-mean-square deviation (RMSE) as the optimization metric, yielding an RMSE of 0.01517 (in the shown example). **D**: The matched spectrum from the WT-compressed dataset is produced. **E:** The scale-matched spectrum is then normalized and compared to the original normalized spectrum, with a computed correlation of 0.99875.

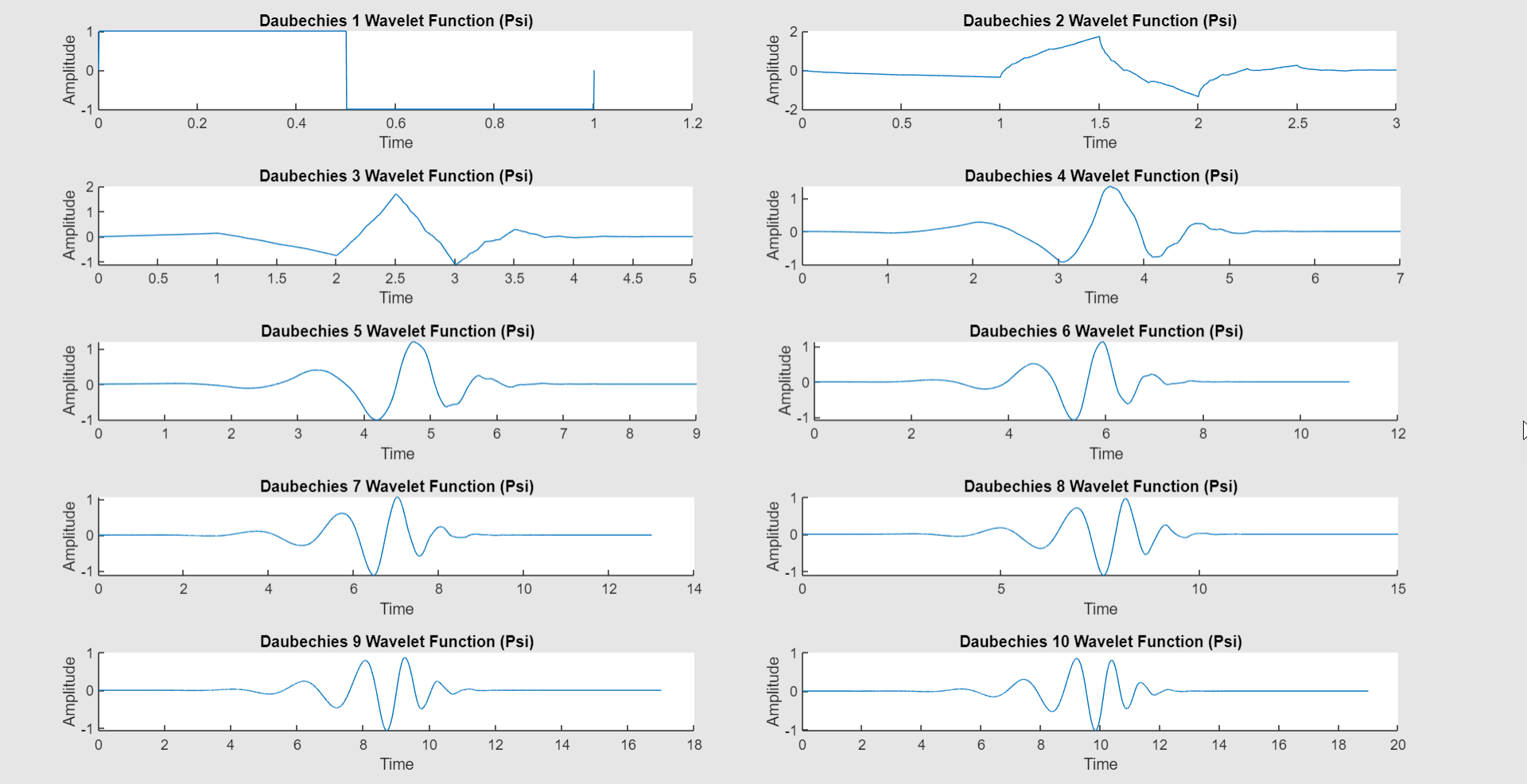

**Figure S3 Wavelets from the *Db* family.** The first 10 members defined by the order number used in this study are shown. *Db3* was the optimal for our dataset.

| DbN_3 | Correlation coefficient |
| --- | --- |
| Db1 | 0.998553 |
| Db2 | 0.999553 |
| Db3 | 0.99948 |
| Db4 | 0.99882 |
| Db5 | 0.999053 |
| Db6 | 0.998577 |
| Db7 | 0.998037 |
| Db8 | 0.997543 |
| Db9 | 0.99707 |
| Db10 | 0.996093 |

**Figure S4 Correlation between wavelet-transformed (WT) compressed spectra and the original spectrum at different orders of *Db.*** The compressed spectra were generated using different Daubechies wavelets with a decomposition level of 3 (*DbN_3,* where *N* is a variable).

Intensity

| Wavelength, nm | original | after db1_1 | after db1_2 | after db1_3 | after db1_4 |
| --- | --- | --- | --- | --- | --- |
| 950.692 | 26820 | 38168.9 | 54495.5 | 78797.9 | 114620 |
| 952.867 | 27112 | 39028.8 | 56967 | 83593.1 | 122019 |
| 955.041 | 27302 | 40073.2 | 58799.5 | 85658.6 | 127236 |
| 957.216 | 27715 | 40835.4 | 59579.5 | 87356.3 | 133042 |
| 959.39 | 28044 | 41526.3 | 60119.5 | 89199 | 132232 |
| 961.565 | 28423 | 41985.2 | 60850 | 91279.7 | 126833 |
| 963.74 | 28641 | 42293.5 | 61404.5 | 93256.1 | 128838 |
| 965.915 | 28837 | 42635.7 | 61970 | 95051.1 | 132096 |
| 968.09 | 29248 | 42743.9 | 62628 | 96443.7 | 132524 |
| 970.265 | 29360 | 42980.8 | 63372.5 | 97419.9 | 127752 |
| 972.44 | 29350 | 43055.7 | 64070.5 | 95994 | 119527 |
| 974.616 | 29501 | 43297.6 | 64850.5 | 91624.8 | 111570 |
| 976.791 | 29563 | 43537.3 | 65536 | 90130.3 | 107291 |
| 978.966 | 29735 | 43704.9 | 66238 | 89917.5 | 107038 |
| 981.142 | 29720 | 43951.6 | 66842.5 | 90589.6 | 107291 |
| 983.318 | 29876 | 44211.9 | 67494 | 91762.7 | 107968 |
| 985.493 | 30055 | 44424.7 | 67102.5 | 93334.6 | 108976 |
| 987.669 | 30144 | 44505.3 | 65168 | 94258.4 | 109890 |
| 989.845 | 30086 | 44863.8 | 64010.5 | 94502.7 | 110290 |
| 992.021 | 30197 | 44959.3 | 63440 | 93938.1 | 109264 |
| 994.197 | 30275 | 45362.3 | 63470 | 92409 | 106228 |
| 996.373 | 30461 | 45440.1 | 63402 | 89664.7 | -1222.5 |
| 998.549 | 30365 | 45761.1 | 63479 | 86612.8 | -1504 |
| 1000.73 | 30533 | 45908.2 | 63630 | 83974.6 | -1380.75 |
| 1002.9 | 30567 | 46244.1 | 63995 | 81152.2 | -678 |
| 1005.08 | 30610 | 46454.1 | 64422.5 | 77769 | 3269.25 |
| 1007.25 | 30579 | 46765.2 | 65072 | 76279.1 | 27.75 |
| 1009.43 | 30752 | 46918.7 | 65453 | 75970.8 | -898 |
| 1011.61 | 30858 | 47133.6 | 65874.5 | 75913.2 | -754.5 |
| 1013.78 | 30763 | 47439.1 | 66422 | 75857.4 | 392 |
| 1015.96 | 31025 | 47605.3 | 66528 | 76036.6 | 2108 |
| 1018.14 | 31084 | 47880.3 | 66649.5 | 76326.5 | 1754.25 |
| 1020.32 | 31098 | 48028.1 | 66586 | 76802.4 | 2289.25 |
| 1022.49 | 31341 | 48156.8 | 66498.5 | 77145.7 | 173.75 |
| 1024.67 | 31200 | 48255.1 | 66062.5 | 77593.7 | -1 |
| 1026.85 | 31411 | 48319.4 | 65537 | 77822.4 | -1.5 |
| 1029.03 | 31303 | 47806.1 | 64726 | 78226.9 | -234.25 |
| 1031.2 | 31505 | 47338.7 | 63623 | 78472.6 | -360 |
| 1033.38 | 31634 | 46631.6 | 62572 | 78740.9 | -132.5 |
| 1035.56 | 31718 | 46319 | 61480 | 78674.8 | -83.25 |
| 1037.74 | 31872 | 46272.4 | 60537.5 | 78322.7 | 742.5 |
| 1039.91 | 31851 | 45857.3 | 59589 | 77357.5 | 2.29017e-12 |
| 1042.09 | 32000 | 45904 | 58784 | 75304.8 | -1785.44 |
| 1044.27 | 32112 | 45887.7 | 57867.5 | -731.502 | -674.226 |
| 1046.45 | 32179 | 45868.6 | 56502.5 | -333.401 | -483.307 |
| 1048.63 | 32210 | 45956.3 | 55256 | -432.396 | -578.06 |
| 1050.81 | 32277 | 45844.6 | 54395.5 | -451.488 | -469.165 |
| 1052.98 | 32481 | 45984.6 | 53910.5 | -398.808 | -458.559 |
| 1055.16 | 32550 | 46065.9 | 53600 | -496.389 | -570.282 |
| 1057.34 | 32548 | 46232.8 | 53492.5 | -498.864 | -366.988 |
| 1059.52 | 32550 | 46246.9 | 53512.5 | -119.501 | -428.153 |
| 1061.7 | 32813 | 46370.6 | 53333 | -166.524 | -76.014 |
| 1063.88 | 32832 | 46565.8 | 53408 | 975.807 | 1057.48 |
| 1066.06 | 32995 | 46852.9 | 53456 | 850.649 | 727.613 |
| 1068.23 | 32989 | 46879.8 | 53399.5 | 114.905 | 94.7523 |
| 1070.41 | 33124 | 47174.6 | 53464.5 | -50.5581 | -54.0937 |
| 1072.59 | 33118 | 47291.3 | 53570 | -160.513 | -264.104 |
| 1074.77 | 33299 | 47351.4 | 53773.5 | -319.612 | -420.375 |
| 1076.95 | 33297 | 47779.2 | 53838.5 | -387.495 | -351.432 |
| 1079.13 | 33470 | 47717 | 53942.5 | -112.076 | -73.5391 |
| 1081.31 | 33443 | 47881 | 54078 | 65.7609 | -24.7487 |
| 1083.49 | 33556 | 47978.6 | 54265 | 186.323 | 220.617 |
| 1085.67 | 33581 | 48271.4 | 54420.5 | 608.112 | 544.826 |
| 1087.85 | 33794 | 48271.4 | 54492 | 729.027 | 767.564 |
| 1090.03 | 33711 | 48297.5 | 54674 | 651.952 | 690.49 |
| 1092.21 | 33936 | 48289 | 54875.5 | 533.866 | 675.287 |
| 1094.39 | 33862 | 48232.5 | 54997 | 1038.39 | 938.331 |
| 1096.57 | 33984 | 48071.2 | 55174 | 597.505 | 544.119 |
| 1098.75 | 33813 | 48099.5 | 55253 | 72.1249 | 129.754 |
| 1100.93 | 34231 | 48130.6 | 55260 | 82.0244 | -21.9203 |
| 1103.11 | 34017 | 48006.2 | 55478.5 | 41.3657 | 59.7505 |
| 1105.29 | 34186 | 47822.3 | 55407 | -99.3485 | 10.253 |
| 1107.47 | 34236 | 47626.5 | 55558.5 | -84.8528 | -79.5495 |
| 1109.65 | 34176 | 47456.1 | 55535.5 | -65.0538 | -83.085 |
| 1111.83 | 34245 | 47130.8 | 55394.5 | -91.9239 | -115.612 |
| 1114.01 | 34313 | 46838 | 55371 | -119.855 | -10.6066 |
| 1116.19 | 34353 | 46478.1 | 55215 | 8.48528 | -147.785 |
| 1118.37 | 34418 | 46041.1 | 54866.5 | -83.085 | -75.6604 |
| 1120.56 | 34368 | 45550.4 | 54387.5 | -49.851 | 19.0919 |
| 1122.74 | 34441 | 45240.7 | 53412.5 | -179.605 | 57.9828 |
| 1124.92 | 34319 | 44894.2 | 52814 | -32.1734 | -32.1734 |
| 1127.1 | 34478 | 44619.9 | -529 | 83.4386 | 77.0746 |
| 1129.28 | 34328 | 44129.8 | -289.5 | 190.919 | 220.264 |
| 1131.46 | 34077 | 43818.7 | -192.5 | 438.406 | 360.978 |
| 1133.64 | 33969 | 43509 | -161.5 | 383.252 | 326.33 |
| 1135.82 | 33672 | 43212 | -33 | -675.5 | -677.5 |
| 1138.01 | 33487 | 42758 | -257.5 | -507.5 | -550.5 |
| 1140.19 | 33490 | 42580.6 | -134 | -216 | -294 |
| 1142.37 | 33073 | 42425.7 | -177 | -253.5 | -172.5 |
| 1144.55 | 32981 | 41944.2 | -199.5 | -79 | -121.5 |
| 1146.73 | 32647 | 41312 | -267.5 | -255.5 | -203 |
| 1148.92 | 32669 | 40697.5 | -141.5 | -181.5 | -135 |
| 1151.1 | 32413 | 40307.9 | -223 | -167 | -204.5 |
| 1153.28 | 32300 | 39811.5 | -150 | -168 | -113 |
| 1155.46 | 32126 | 39582.4 | -180.5 | -257.5 | -203.5 |
| 1157.64 | 32193 | 39274.1 | 36 | -313.5 | -189.5 |
| 1159.83 | 31975 | 39038.7 | -107 | -137.5 | -296 |
| 1162.01 | 32100 | 38892.3 | -40.5 | -186 | -156.5 |
| 1164.19 | 32108 | 38832.9 | -184 | -120 | -320 |
| 1166.38 | 32112 | 38860.5 | 98 | -63.5 | -112 |
| 1168.56 | 31986 | 38744.5 | 211 | -128 | -97 |
| 1170.74 | 32137 | 38679.4 | 356.5 | -190.5 | -8.5 |
| 1172.92 | 32012 | 38796.1 | 305 | -71.5 | -143 |
| 1175.11 | 32022 | 38788.3 | 181.5 | -25.5 | -198.5 |
| 1177.29 | 31931 | 38744.5 | 27 | -3.14399e-12 | 71 |
| 1179.47 | 32038 | 38677.3 | 63 | 137 | 90.5 |
| 1181.66 | 31937 | 38685.1 | -63 | 458 | 434 |
| 1183.84 | 32061 | 38813.8 | -118 | 346 | 332 |
| 1186.03 | 31982 | 38712 | -130 | 259 | 122 |
| 1188.21 | 31995 | 38769.3 | -178 | -5.55166e-12 | 125.5 |
| 1190.39 | 32066 | 38687.9 | -24.5 | 55.5 | -74.5 |
| 1192.58 | 32206 | 38876 | -153 | -3.5 | -39.5 |
| 1194.76 | 32066 | 38720.5 | -154 | -62 | 65 |
| 1196.94 | 32219 | 38794.7 | -17.5 | -75 | -137 |
| 1199.13 | 32214 | 38889.5 | -103 | -165 | -120.5 |
| 1201.31 | 32237 | 38918.5 | 2 | -115 | -28.5 |
| 1203.5 | 32355 | 39037.2 | -25.5 | -219 | -21 |
| 1205.68 | 32306 | 38973.6 | 150 | -96.5 | -200.5 |
| 1207.87 | 32320 | 38898.7 | 163.5 | -124.5 | -68.5 |
| 1210.05 | 32523 | 39007.5 | 147.5 | -106.5 | -90.5 |
| 1212.23 | 32416 | 39105.8 | 239 | -72 | 123.5 |
| 1214.42 | 32631 | 39181.5 | 282 | -61 | -20.5 |
| 1216.61 | 32570 | 39216.8 | 300 | 73 | 122.5 |
| 1218.79 | 32778 | 39388 | 241 | 46 | 127 |
| 1220.97 | 32689 | 39448.8 | 278 | 60.5 | -1 |
| 1223.16 | 32916 | 39360.4 | 206.5 | 293 | 152 |
| 1225.35 | 32842 | 39427.6 | 244 | 305 | 200.5 |
| 1227.53 | 32969 | 39653.1 | 147 | 161 | 315.5 |
| 1229.71 | 32933 | 39547.1 | 338.5 | 259 | 208 |
| 1231.9 | 33095 | 39540 | 443.5 | 214.5 | 253 |
| 1234.09 | 33124 | 39726.7 | 299 | 152.5 | 313.5 |
| 1236.27 | 33334 | 39768.4 | 189.5 | 165.5 | 185 |
| 1238.46 | 33239 | 39832.7 | 183.5 | 145.5 | 162 |
| 1240.64 | 33307 | 39810.8 | 56 | 234.5 | 242 |
| 1242.83 | 33190 | 39933.1 | 26.5 | 370 | 409 |
| 1245.02 | 33463 | 39872.3 | 4.5 | 123.5 | 277 |
| 1247.2 | 33502 | 39858.2 | 16 | 119.5 | 113.5 |
| 1249.39 | 33555 | 39976.3 | 6 | 74.5 | 34 |
| 1251.57 | 33495 | 40121.2 | -17 | 37.5 | 8.5 |
| 1253.76 | 33680 | 40001 | -50.5 | 43.5 | 33 |
| 1255.94 | 33694 | 40101.4 | -37.5 | -10.5 | -30 |
| 1258.13 | 33694 | 40092.2 | -38 | 73 | 103 |
| 1260.32 | 33585 | 40122.7 | -0.5 | -43.5 | -34.5 |
| 1262.51 | 33633 | 40196.2 | -84.5 | -71 | 189 |
| 1264.69 | 33655 | 40075.3 | 36.5 | -1.5 | -40.5 |
| 1266.88 | 33797 | 40194.8 | -32 | -68 | 40.5 |
| 1269.07 | 33682 | 40145.3 | -52 | -49 | -26 |
| 1271.25 | 33911 | 40127.6 | 33.5 | -11 | -33.5 |
| 1273.44 | 33715 | 40014.5 | -139 | 10 | -60 |
| 1275.63 | 33878 | 40196.2 | -162 | -57.5 | -91 |
| 1277.81 | 33562 | 40021.5 | -82.5 | -19.5 | -3.5 |
| 1280 | 33765 | 40006 | -15 | 0.5 | -37.5 |
| 1282.19 | 33587 | 39861 | -47 | -19 | -60.5 |
| 1284.38 | 33563 | 39914.8 | -8 | -100 | -0.5 |
| 1286.56 | 33464 | 39837 | 36 | -56 | -33.5 |
| 1288.75 | 33585 | 39762.7 | -35.5 | 42 | 10.5 |
| 1290.94 | 33535 | 39592.3 | 66 | 24.5 | 6.5 |
| 1293.13 | 33439 | 39447.4 | -39.5 | -7.5 | -40.5 |
| 1295.31 | 33338 | 39104.4 | -47.5 | 62 | -77.5 |
| 1297.5 | 33423 | 38749.5 | 14.5 | -1.5 | -105.5 |
| 1299.69 | 33406 | 38475.8 | 10 | -109.5 | 14.5 |
| 1301.88 | 33253 | 37999.2 | 104 | 3.5 | -4.5 |
| 1304.07 | 33183 | 37805.5 | 195.5 | -9 | -41 |
| 1306.26 | 33066 | 37585.6 | 219.5 | -1.5 | -93 |
| 1308.44 | 32751 | -252.437 | 217.5 | -58.5 | 4 |
| 1310.63 | 32882 | -406.586 | 122 | -47.5 | -43 |
| 1312.82 | 32644 | -213.546 | -337.997 | 83.5 | 36.5 |
| 1315.01 | 32590 | -226.274 | -215.668 | 78 | 191 |
| 1317.2 | 32358 | -217.082 | -237.588 | 238 | 161.5 |
| 1319.39 | 32327 | -127.279 | -117.38 | 198.5 | 126 |
| 1321.58 | 32166 | -82.0244 | -82.0244 | -4.5 | 141.5 |
| 1323.76 | 32124 | -50.9117 | 37.4767 | -85.5599 | -217.082 |
| 1325.95 | 31942 | -109.602 | -245.366 | -251.73 | -195.161 |
| 1328.14 | 31753 | -144.25 | -59.397 | -156.271 | -310.42 |
| 1330.33 | 31542 | -237.588 | -28.2843 | -147.078 | -271.529 |
| 1332.52 | 31465 | -67.8823 | -89.8026 | -22.6274 | -127.279 |
| 1334.71 | 31297 | -284.964 | -95.4594 | -73.5391 | -48.0833 |
| 1336.9 | 31087 | -69.2965 | -44.5477 | -149.2 | -12.0208 |
| 1339.09 | 31124 | -140.714 | -218.496 | -21.2132 | -63.6396 |
| 1341.28 | 30955 | -123.744 | -22.6274 | -102.53 | -86.9741 |
| 1343.47 | 30826 | -141.421 | -195.161 | -109.602 | -57.9828 |
| 1345.66 | 30622 | -186.676 | -106.066 | -140.007 | -10.6066 |
| 1347.85 | 30438 | -95.4594 | -166.17 | -33.234 | -45.9619 |
| 1350.04 | 30446 | -113.137 | -26.163 | -14.8492 | -125.158 |
| 1352.23 | 30233 | -199.404 | -188.798 | -70.7107 | -146.371 |
| 1354.42 | 30245 | -43.8406 | -74.9533 | -53.033 | -146.371 |
| 1356.61 | 30075 | -63.6396 | -193.04 | 21.9203 | -18.3848 |
| 1358.8 | 29864 | -171.12 | -26.8701 | 2.82843 | 21.9203 |
| 1360.99 | 29916 | -115.258 | 44.5477 | -49.4975 | -108.187 |
| 1363.18 | 29830 | -48.0833 | 31.1127 | 17.6777 | -152.735 |
| 1365.37 | 29713 | -152.735 | 27.5772 | -8.48528 | -60.1041 |
| 1367.56 | 29604 | -86.9741 | -77.0746 | 68.5894 | 8.48528 |
| 1369.75 | 29433 | -126.572 | -32.5269 | -82.0244 | -21.9203 |
| 1371.94 | 29409 | -201.525 | -89.0955 | -10.6066 | -168.291 |
| 1374.13 | 29187 | -258.801 | -192.333 | -94.7523 | -77.7817 |
| 1376.33 | 28912 | -71.4178 | 14.8492 | -65.7609 | -20.5061 |
| 1378.52 | 28774 | -128.693 | -79.196 | -54.4472 | -84.8528 |
| 1380.71 | 28531 | -207.889 | 69.2965 | -10.6066 | -125.865 |
| 1382.9 | 28384 | -101.116 | -20.5061 | 84.1457 | 2.82843 |
| 1385.09 | 28208 | -235.467 | 7.77817 | 0.707107 | -137.179 |

**Figure S5 Changes of the representative spectrum after the WT application**. *Db1* WT was applied with different level of decomposition. The spectra were recorded from the same ROI. Higher levels of decomposition produced “compressed spectrum” with higher level of intensities.

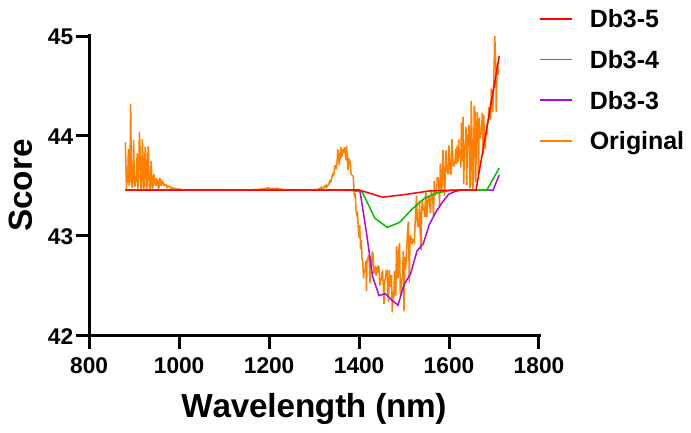

**Figure S6** **Quality score of the original and WT applied datasets as calculated by BRISQUE algorithm. The** method calculates the perceptual quality score of the image by using an SVM model trained on an image database. Scale from 0 (ideal quality) to 100 (worst quality). Lower level (higher score) of image quality below 950 and above 1650 nm is a result of low sensitivity of the InGaAs sensor, that can be partially improved by WT.

| **A** |  |
| --- | --- |
| 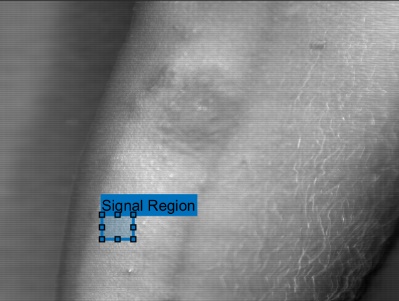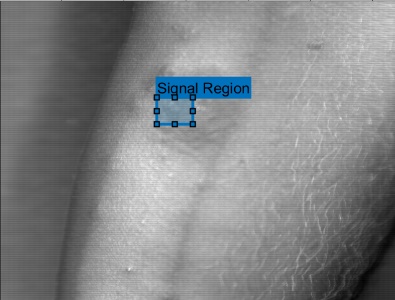 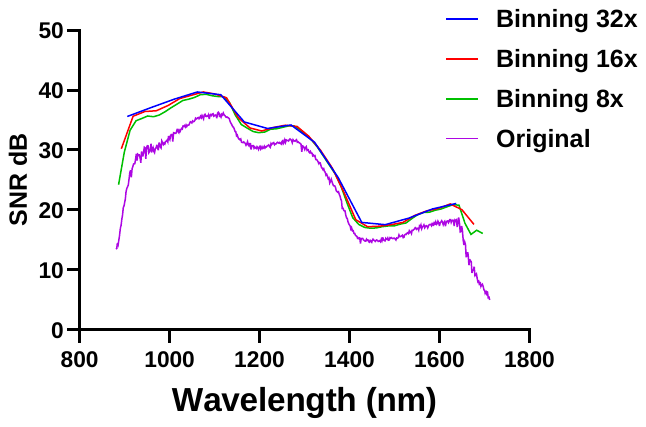 | |
| **B** | **C** |
| 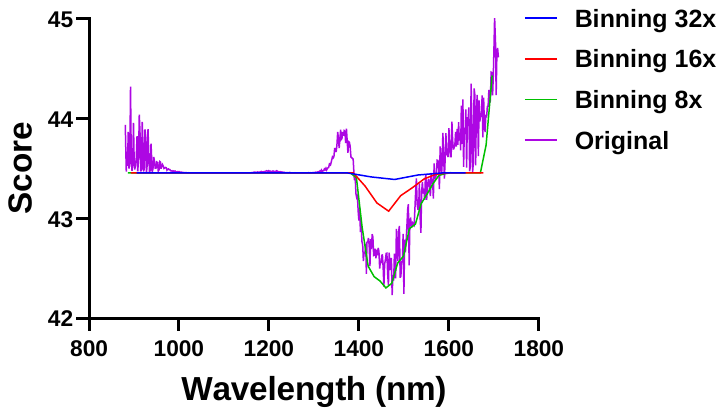 | 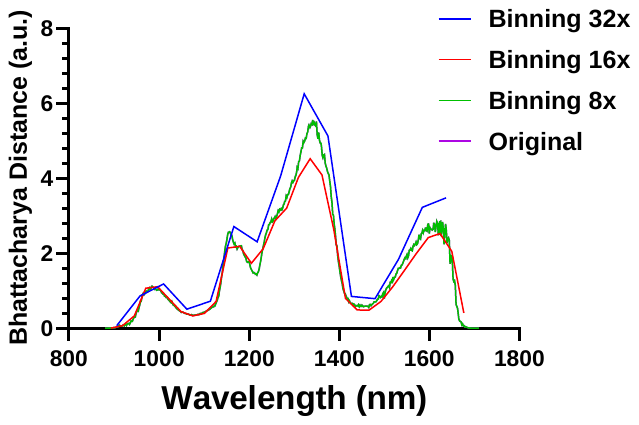 |

**Figure S****7 Metrics of the images after spectral binning**. **A**: SNR from the areas selected from the tissue (signal) and background (noise) **B**: Quality score of each frame using BRISQUE, **C:** Contrast between the lesion and healthy tissue using Bhattacharya distance method.
